# Increased LPS levels coexist with systemic inflammation and result in monocyte activation in severe COVID-19 patients

**DOI:** 10.1101/2021.06.24.21259468

**Authors:** Paula C Teixeira, Gilson P Dorneles, Paulo C Santana Filho, Igor M da Silva, Lucas L Schipper, Isabelle AL Postiga, Carla Andretta Moreira Neves, Luiz Carlos Rodrigues Júnior, Alessandra Peres, Janeusa Trindade de Souto, Simone Gonçalves da Fonseca, Sarah Eller, Tiago F Oliveira, Liane N. Rotta, Claudia Thompson, Pedro R T Romão

## Abstract

This study aimed to evaluate the link between microbial translocation markers and systemic inflammation at the earliest time-point after hospitalization and at the last 72 h of hospitalization in survivors and non-survivors COVID-19 patients. Sixty-six SARS-CoV-2 RT-PCR+ infected patients and nine non-COVID-19 pneumonia controls were admitted in this study. Blood samples were collected at hospital admission (T1) (Controls and COVID-19+ patients) and 0-72 h before hospital discharge (T2, alive or dead) to analyze systemic cytokines and chemokines, LPS concentrations and soluble CD14 (sCD14) levels. THP-1 human monocytic cell line was incubated with plasma from survivors and non-survivors COVID-19 patients and their phenotype, activation status, TLR4, and chemokine receptors were analyzed by flow cytometry. COVID-19 patients presented higher IL-6, IFN-γ, TNF-α, TGF-β1, CCL2/MCP-1, CCL4/MIP-1β, and CCL5/RANTES levels than controls. Moreover, LPS and sCD14 were higher at hospital admission in SARS-CoV-2-infected patients. Non-survivors COVID-19 patients had increased LPS levels concomitant with higher IL-6, TNF-α, CCL2/MCP-1, and CCL5/RANTES levels at T2. Increased expression of CD16 and CCR5 were identified in THP-1 cells incubated with the plasma of survivor patients obtained at T2. The incubation of THP-1 with T2 plasma of non-survivors COVID-19 leads to higher TLR4, CCR2, CCR5, CCR7, and CD69 expression. In conclusion, increased microbial translocation during hospitalization coexist with the inflammatory condition of SARS-CoV-2 infection and could lead to higher monocyte activation in non-survivors COVID-19 patients.

## Introduction

Severe acute respiratory syndrome coronavirus 2 (SARS-CoV-2) infection causes Coronavirus disease 2019 (COVID-19), a disease with diverse clinical manifestations (1). While most COVID-19 cases are asymptomatic or mild, severe form of COVID-19 can occur with detrimental manifestations such as acute respiratory distress syndrome (ARDS), multi-organ failure, and death (2). The COVID-19 immunopathology is mainly characterized by a hyperinflammatory state, strong innate cells response, and lymphocyte activation with an exhausted profile (3). Furthermore, there is a clear association between COVID-19 severity and higher levels of systemic cytokines (4).

The mechanism underlying the inflammatory state of COVID-19 is not fully understood at this time. Although the respiratory tract is the main site of infection for COVID-19, the pathogenesis of the disease can also involve different organs and systems, such as the heart, kidneys, intestines, vasculature, and liver (3). In the gastrointestinal (GI) tract, symptoms such as diarrhea and abdominal distention have been reported in several patients (5). Furthermore, enterocytes in the ileum and colon express the angiotensin-converting enzyme 2 (ACE-2) receptor and may serve as a site for SARS-CoV-2 entrance and predispose to enteric infection (6). Additionally, RNA viral particles were detected in stool samples, arising the possibility that bacterial translocation and microbial products from the GI tract to the peripheral blood might contribute to the hyperinflammatory state and COVID-19 severity (7).

Moreover, systemic inflammation caused by lung infection can lead to the disruption of the gut barrier integrity and increase the permeability to gut microbes and microbial products (8–10). Interestingly, disruption in gut barriers and microbial translocation is more likely to occur in older individuals and individuals with chronic diseases, such as type 2 diabetes, cardiovascular diseases, and obesity, which contribute to the exacerbation of systemic inflammation by activating innate immune cells, mainly monocytes and neutrophils (8).

Lipopolysaccharide (LPS) is a product derived from the membrane of gram-negative bacteria that acts as a potent immune-activating stimulus when recognized by innate immune cells expressing toll-like receptor 4 (TLR4) (11). Then, innate immune cells become activated and produce high amounts of proinflammatory mediators including cytokines, such as interleukin (IL)-6 and tumor necrosis factor-alpha (TNF-α) and chemokines (i.e., CCL2/MCP-1 and CCL5/RANTES) (12). In this sense, peripheral blood LPS is a well-recognized marker of translocation of microorganisms after events that compromise the integrity of the intestinal mucosa (13,14). Besides, soluble CD14 (sCD14) is released after cleavage from the membrane form (mCD14) on the surface of monocytes through exposure to LPS stimulation and is considered a marker of microbial translocation (14,15).

Emerging evidence indicates that severe SARS-CoV-2 infection induces disruption in the gut barrier contributing to the systemic spread of bacteria and microbial products which affect the host’s response to the infection (9,16,17). However, the time course of the presence of microbial product in the peripheral blood of COVID-19 patients during hospitalization remains unknown. Here, we investigated the link between microbial translocation markers and systemic inflammation in the hospital admission and at the discharge time from the hospital in survivors and non-survivors COVID-19 patients. We hypothesized that microbial translocation markers are associated with systemic inflammation and could contribute to the death of hospitalized COVID-19 patients.

## Methods

### Study population

Hospitalized COVID-19 patients with confirmed positive SARS‐CoV‐2 RT-PCR test on nasopharyngeal specimens, were consecutively recruited after admission in the COVID-19 Unit of Hospital São Camilo (Esteio/RS, Brazil) between June/2020 and December/2020 to a clinical cohort study. The study was approved by the UFCSPA Ethics Committee (CAAE: 38886220.0.0000). Informed consent was obtained from those legally responsible for the patients after the procedures had been explained. The authors signed an agreement to preserve patient and staff anonymity related to the use of this data. All patients were confirmed to COVID-19 infection according to the World Health Organization interim guidance and Diagnosis and Treatment Guideline for Novel Coronavirus Pneumonia and were positive for at least two nucleic acid tests for SARS-CoV-2 (18).

Clinical and socio-demographic data were collected from the patient’s electronic medical records upon admission to the unit. Body mass index was calculated from weight and height data and was classified according to age. Blood samples from 9 age-and sex-matched SARS-CoV-2 RT-PCR negative controls admitted to hospital with pneumonia were also obtained.

### Blood collection and Laboratory Analysis

Blood samples were collected from the antecubital vein of the patients into 4 mL tubes with EDTA as anti-coagulant. Blood collection were taken at the earliest time-point after hospitalization (T1) and before discharge time from the hospital (T2: 0 to 72 hours before leaving hospital or death). Plasma samples were obtained by centrifugation (2000g, 10 minutes), aliquoted and immediately kept at -80ºC until analysis.

Plasma levels of IL-6, IL-10, TNF-α, TGF-β1 (all from Invitrogen Life Sciences, USA), IFN-γ, CCL2/MCP-1, CCL4/MIP-1β, and CCL5/RANTES (all from Peprotech, USA) were analyzed by *Enzyme Linked Immunosorbent Assay* (ELISA) following the manufacturer’s procedure using a microplate reader (EzBiochrom, USA). The intra-assay coefficient of variability was < 7.5%. The detection limits of each cytokine were: IL-6, 2 – 200 pg/mL; IL-10, 4 – 200 pg/mL; IFN-γ, 10 – 300 pg/mL; TNF-α, 2 – 200 pg/mL; TGF-β1, 2 – 500 pg/mL; CCL5/RANTES, 20 – 1000 pg/mL; CCL4/MIP-1β, 50 – 1500 pg/mL; CCL2/MCP-1, 50 – 1000 pg/mL. The soluble form of CD14 was analyzed using Human CD14 ELISA Kit from Invitrogen (USA), with detection limits ranging 8.23 – 8000 pg/mL.

### Liquid chromatography-tandem mass spectrometry determination of LPS

LPS concentration was determined by quantitation of 3-hydroxytetradecanoic acid as described by Pais de Barros and colleagues (19). Briefly, 150 µL of plasma was hydrolyzed with 75 µL of NaCl 150 mM, 300 µL of HCl 8 mol for 4 h at 90 °C and extracted with 4 mL of a hexane. Samples were dried under a nitrogen stream and resuspended in 50 μL of methanol immediately prior to the injection in the analytical system. A Nexera UFLC system coupled to a LCMS-8040 triple quadrupole mass spectrometer (Shimadzu, Kyoto, Japan) was used for the analysis. The electrospray parameters were set in the negative ion mode ([M-H]-) as follows: capillary voltage, 3000 V; desolvation line temperature, 250 ºC; heating block temperature, 500 ºC; drying gas, 18/L min; and nebulizing gas, 2 L/min. Collision-induced dissociation was obtained with 230 kPa argon pressure. Analyses were carried out with multiple reaction monitoring (MRM) by using the following fragmentation: m/z 243.1 → m/z 59.0. The chromatographic separation was conducted with an Acquity UPLC® C18 column (2.1 x 50 mm, 1.7 μm particle size) (Waters Corporation, Ireland). The analyses were performed in gradient elution mode with a flow rate of 0.4 mL/min and the gradient mobile phase system consisted of water (solvent A) and acetonitrile (solvent B) both fortified with 0.2 % acetic acid as follow: 0 – 1.5 min, 75 – 100% of B; 1.5 – 2.0 min, 100% of B; 2.0 – 2.1 min, 100 – 75 % of B; 2.1 – 5.5 min, 75 % B. The column oven was kept at 50 °C. The data were processed using LabSolutions software (Shimadzu, Kyoto, Japan). Calibration curves were constructed at intervals of 0.2 -1000 ng/mL.

### Cell Culture and Flow cytometry

THP-1 human monocyte cells (ATCC TIB-202) (2 x 10^5^ cells/well) were cultured in RPMI media (Sigma-Aldrich, USA) supplemented with 10% fetal bovine serum (FBS, Gibco, USA) and 1% penicillin and streptomycin (both from Sigma-Aldrich) in 96 well plates for 24 hours. Afterwards, the media plus FBS were removed; cells were washed with phosphate-saline buffer and incubated with RPMI supplemented with 10% serum of survivors and non-survivors COVID-19 patients for 15 hours. Then, cells were labelled with monoclonal antibodies (all anti-human) conjugated with specific fluorochromes: CD14 Fitc, CD16 Pe, TLR4 Pe, CD69 Percp-Cy5.5, CD192 (CCR2) Pe, CD195 (CCR5), CD197 (CCR7) Fitc, HLA-DR PercP-Cy5.5 (all from EbioScience, ThermoFisher, USA). 10.000 events were acquired using CELLQuest Pro Software (BD Bioscience, USA) on a FACSCalibur flow cytometer (BD Bioscience, USA) to determine cell phenotype. THP-1 were identified and gated according to each forward scatter (FSC) and side scatter (SSC) profile and the mean fluorescence intensity (MFI) was evaluated.

### Statistical Analysis

Normality of data was checked by Kolmogorov‐Smirnov, and the values were presented as mean ± 95%CI for demographic and clinical characteristics, and mean ± standard deviation (SD) for inflammatory mediators. Participant’s characteristics and inflammatory analysis were between-groups compared through a one‐way ANOVA followed by Bonferroni’s post‐hoc for multiple comparisons. The time-course of inflammatory response and immune variables was analyzed by a generalized linear mixed model. Correlation analyses between markers microbial translocation and inflammatory mediators were performed using Pearson’s Coefficient of Correlation. P values ≤ 0.05 were considered statistically significant. The SPSS 22.0 (IBM Inc, EUA) software was used in all analysis.

## Results

### Characteristics of the study population

Firstly, we analyzed the baseline characteristics and inflammatory mediators in controls and COVID-19 hospitalized patients. To this, 66 patients admitted at COVID-19 care unit due to COVID-19 complications were enrolled in this study. Additionally, 9 COVID-19-controls were analyzed. COVID-19 patients presented higher body mass and body mass index than controls (p<0.01), and 53.8% of patients were classified as obese. The mean length of stay in the hospital were 21.1 (15.6 – 26.6) days for COVID-19 patients, 84.8% patients were admitted to the intensive care unit (ICU) due to COVID-19 complications, and 34.8% patients died due to complications of SARS-CoV-2 infection. Forty-three (43) patients survived to SARS-CoV-2 infection and were classified as COVID-19 survivors. Patients and control data are shown in Table 1.

**Table 1.**
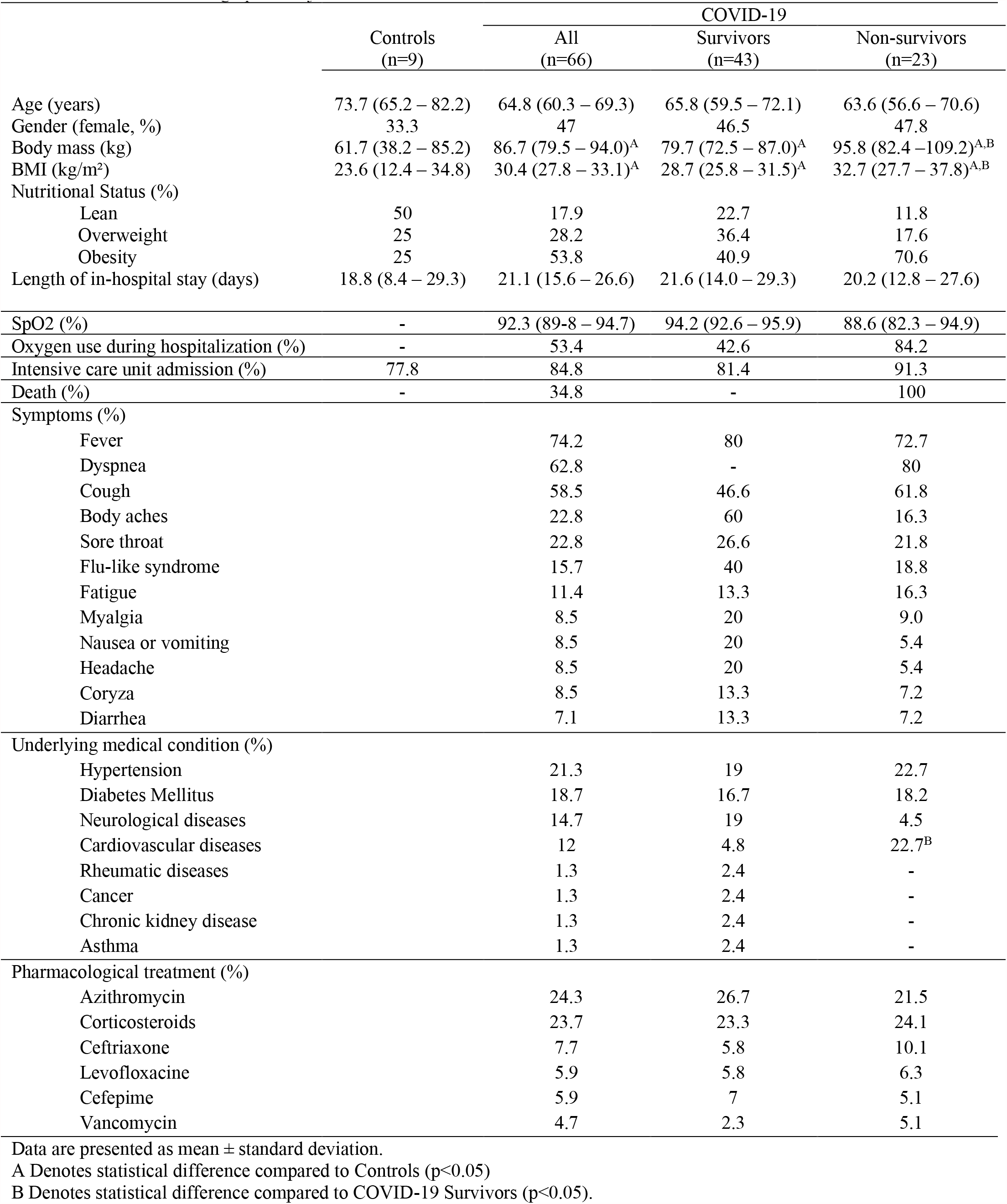
Clinical and demographic subject’s characteristics

|  | Controls<br>(n=9) | COVID-19 |  |  |
| --- | --- | --- | --- | --- |
|  |  | All<br>(n=66) | Survivors<br>(n=43) | Non-survivors<br>(n=23) |
| Age (years) | 73.7 (65.2 – 82.2) | 64.8 (60.3 – 69.3) | 65.8 (59.5 – 72.1) | 63.6 (56.6 – 70.6) |
| Gender (female, %) | 33.3 | 47 | 46.5 | 47.8 |
| Body mass (kg) | 61.7 (38.2 – 85.2) | 86.7 (79.5 – 94.0)<sup>A</sup> | 79.7 (72.5 – 87.0)<sup>A</sup> | 95.8 (82.4 – 109.2)<sup>A,B</sup> |
| BMI (kg/m<sup>2</sup>) | 23.6 (12.4 – 34.8) | 30.4 (27.8 – 33.1)<sup>A</sup> | 28.7 (25.8 – 31.5)<sup>A</sup> | 32.7 (27.7 – 37.8)<sup>A,B</sup> |
| Nutritional Status (%) |  |  |  |  |
| Lean | 50 | 17.9 | 22.7 | 11.8 |
| Overweight | 25 | 28.2 | 36.4 | 17.6 |
| Obesity | 25 | 53.8 | 40.9 | 70.6 |
| Length of in-hospital stay (days) | 18.8 (8.4 – 29.3) | 21.1 (15.6 – 26.6) | 21.6 (14.0 – 29.3) | 20.2 (12.8 – 27.6) |
| SpO2 (%) | - | 92.3 (89.8 – 94.7) | 94.2 (92.6 – 95.9) | 88.6 (82.3 – 94.9) |
| Oxygen use during hospitalization (%) | - | 53.4 | 42.6 | 84.2 |
| Intensive care unit admission (%) | 77.8 | 84.8 | 81.4 | 91.3 |
| Death (%) | - | 34.8 | - | 100 |
| Symptoms (%) |  |  |  |  |
| Fever |  | 74.2 | 80 | 72.7 |
| Dyspnea |  | 62.8 | - | 80 |
| Cough |  | 58.5 | 46.6 | 61.8 |
| Body aches |  | 22.8 | 60 | 16.3 |
| Sore throat |  | 22.8 | 26.6 | 21.8 |
| Flu-like syndrome |  | 15.7 | 40 | 18.8 |
| Fatigue |  | 11.4 | 13.3 | 16.3 |
| Myalgia |  | 8.5 | 20 | 9.0 |
| Nausea or vomiting |  | 8.5 | 20 | 5.4 |
| Headache |  | 8.5 | 20 | 5.4 |
| Coryza |  | 8.5 | 13.3 | 7.2 |
| Diarrhea |  | 7.1 | 13.3 | 7.2 |
| Underlying medical condition (%) |  |  |  |  |
| Hypertension |  | 21.3 | 19 | 22.7 |
| Diabetes Mellitus |  | 18.7 | 16.7 | 18.2 |
| Neurological diseases |  | 14.7 | 19 | 4.5 |
| Cardiovascular diseases |  | 12 | 4.8 | 22.7<sup>B</sup> |
| Rheumatic diseases |  | 1.3 | 2.4 | - |
| Cancer |  | 1.3 | 2.4 | - |
| Chronic kidney disease |  | 1.3 | 2.4 | - |
| Asthma |  | 1.3 | 2.4 | - |
| Pharmacological treatment (%) |  |  |  |  |
| Azithromycin |  | 24.3 | 26.7 | 21.5 |
| Corticosteroids |  | 23.7 | 23.3 | 24.1 |
| Ceftriaxone |  | 7.7 | 5.8 | 10.1 |
| Levofloxacin |  | 5.9 | 5.8 | 6.3 |
| Cefepime |  | 5.9 | 7 | 5.1 |
| Vancomycin |  | 4.7 | 2.3 | 5.1 |

Data are presented as mean ± standard deviation.
A Denotes statistical difference compared to Controls (p<0.05)
B Denotes statistical difference compared to COVID-19 Survivors (p<0.05).

Interestingly, COVID-19 non-survivors (n=23, 30.7% of COVID-19 patients) were heavier than controls (body mass, p=0.01; BMI, p=0.01) and COVID-19 survivors (n=43, 57.3% COVID-19 patients) (body mass, p=0.03; BMI, p=0.02), but no difference between groups was found in the hospital length of stay (p>0.05). COVID-19 non-survivors required more oxygen use during hospitalization than COVID-19 survivors (p=0.002), but similar rates of ICU admission were observed (p>0.05). The most reported symptoms by COVID-19 patients were fever (74.2%), dyspnea (62.8%), and cough (58.5%). Furthermore, 21.7% of COVID-19 patients were diagnosed with hypertension, 18.7% with diabetes mellitus 2, 14.7% with some neurologic disease and 12% with some cardiovascular disease. Regarding pharmacological treatment, azithromycin (24.3%) and corticosteroids therapy (23.7%) were the predominant medicine adopted by medical staff during hospitalization. There was no difference between groups regarding symptoms, comorbidities, or pharmacological therapy during hospitalization (p<0.05), unless the prevalence of cardiovascular diseases (COVID-19 survivors, 4.8; COVID-19 non-survivors, 22.7%; p=0.04).

### Systemic inflammatory mediators in survivors and non-survivors COVID-19 patients

COVID-19 patients presented a higher inflammatory profile at hospital admission compared to controls as presented in Table 2. COVID-19 patients had increased systemic levels of IL-6 (p<0.01), IFN-γ (p<0.01), TNF-α (p<0.01), CCL5/RANTES (p<0.01), CCL4/MIP-1β (p<0.01) and CCL2/MCP-1 (p<0.01) but diminished circulating levels of TGF-β1 (p<0.05). When COVID-19 patients were stratified in survivors and non-survivors, both groups presented higher IL-6 (p<0.01), CCL2/MCP-1 (p<0.01) and CCL4/MIP-1β (p<0.01) levels at hospital admission compared to controls. However, only non-survivors had increased IFN-γ (p<0.01) and TNF-α (p<0.05) compared to controls, and higher TNF-α levels (p<0.05) than survivors. Conversely, survivors had higher levels of CCL5/RANTES (p<0.01) and CCL4/MIP-1β (p<0.05) compared to control and non-survivors groups, respectively. Furthermore, diminished TGF-β1 levels were found in both survivors and non-survivors COVID-19 patients compared to controls (p<0.01). Finally, a strong monocytopenia was observed in COVID-19 non-survivors compared to COVID-19 survivors (p=0.01). These data indicate that COVID-19 patients have higher levels of inflammatory cytokines at the hospital admission, and higher TNF-α plasma levels that may be associated with death.

**Table 2.**
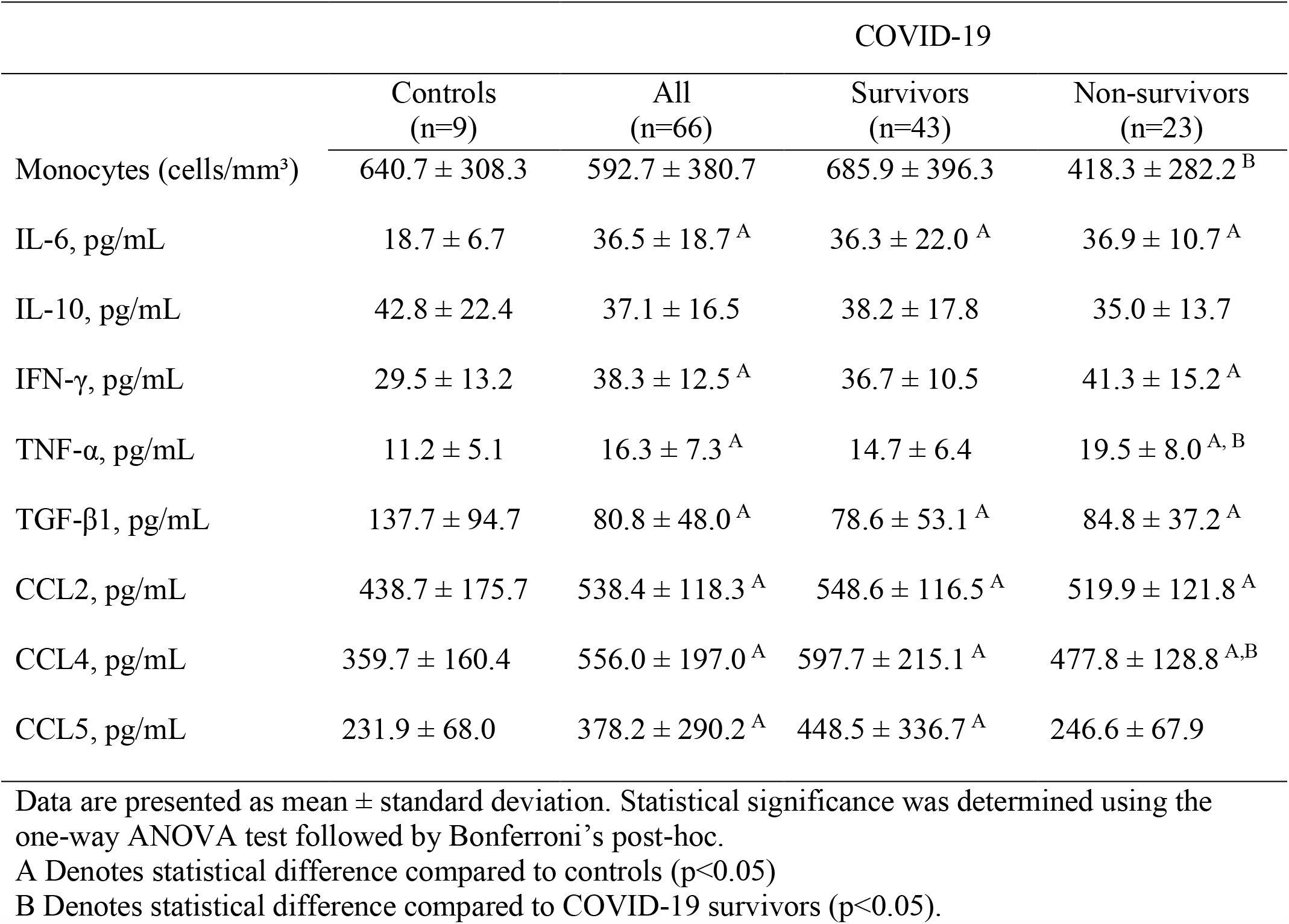
Systemic inflammatory mediators in survivors and non-survivors COVID-19 hospitalized patients.

### Increased of microbial translocation markers in plasma of COVID patients are associated with systemic inflammation

Markers of microbial translocation were found in the peripheral blood of COVID-19 hospitalized patients. In fact, COVID-19 subjects, independently of the outcome group, presented higher sCD14s (p<0.001 for all comparisons) and LPS (controls vs. COVID-19 patients, p<0.001; controls vs. COVID-19 survivors, p<0.001; controls vs. COVID-19 non-survivors, p<0.05) levels at time of hospital admission (Figure 1). We examined the Pearson’s Coefficient Correlation between inflammatory mediators, sCD14 and LPS for each participant. We highlight the significant correlations of sCD14 with TGF-β (r= -0.46; p<0.001), CCL4/MIP-1β (r= 0.46; p<0.001) and CCL2/MCP-1 (r= 0.29; p=0.03). LPS was correlated with TNF-α (r= -0.32; p<0.004), CCL5/RANTES (r= 0.49; p<0.001) and CCL4/MIP-1β (r= 0.27; p=0.01) (Figure 2).

**Figure 1.**
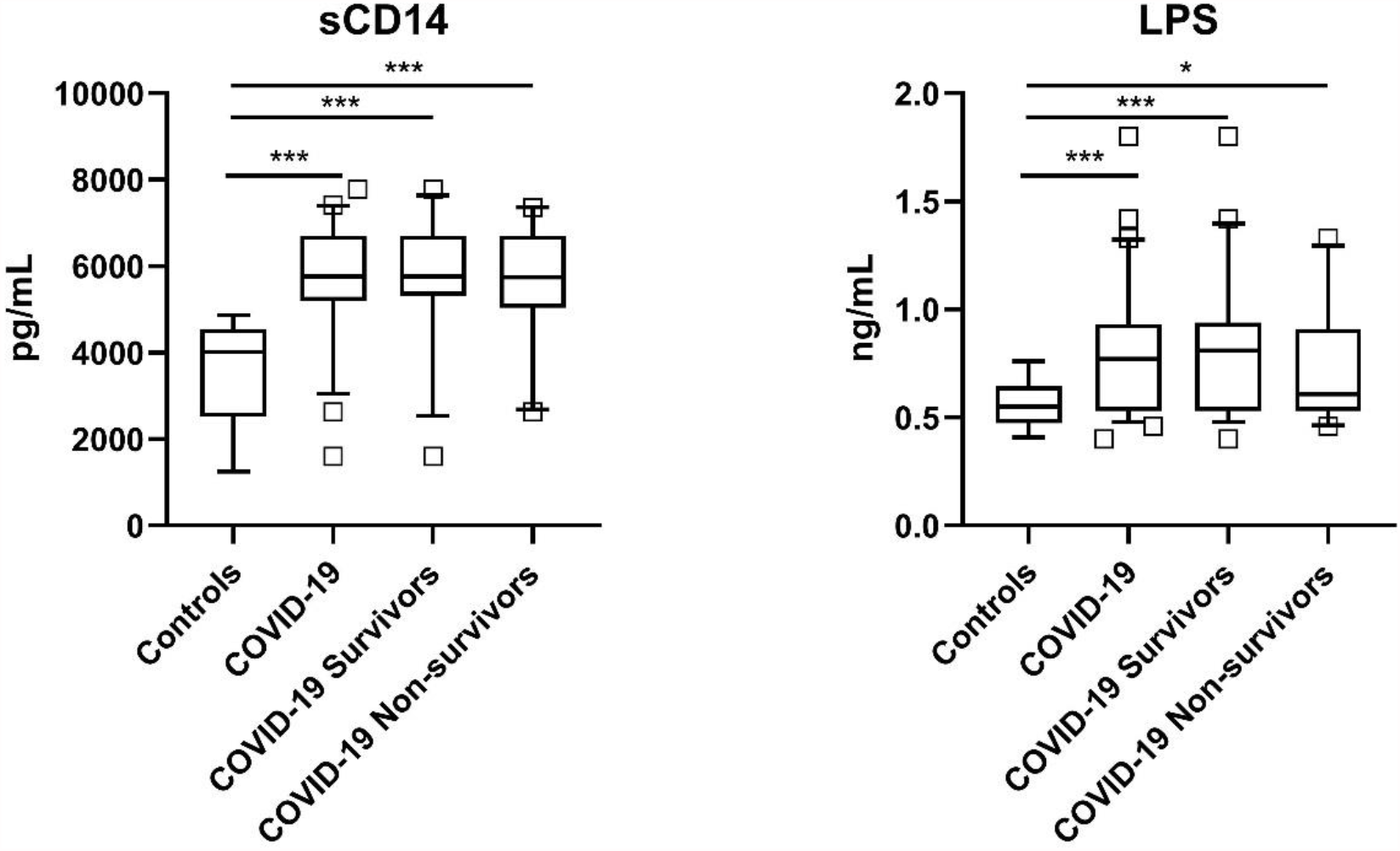
Microbial translocation markers in plasma of COVID-19 patients at time at hospital admission. The systemic levels of soluble CD14 (A) and LPS (B) in controls (n=9) and COVID-19 survivors (n=43) and non-survivors (n=23) patients were compared using one-way ANOVA followed by Bonferroni’s post-hoc for multiple comparisons (p<0.05). Box with 5-95 percentile data were used to show each data. * Indicates p<0.05. *** Indicates p<0.001

**Figure 2.**
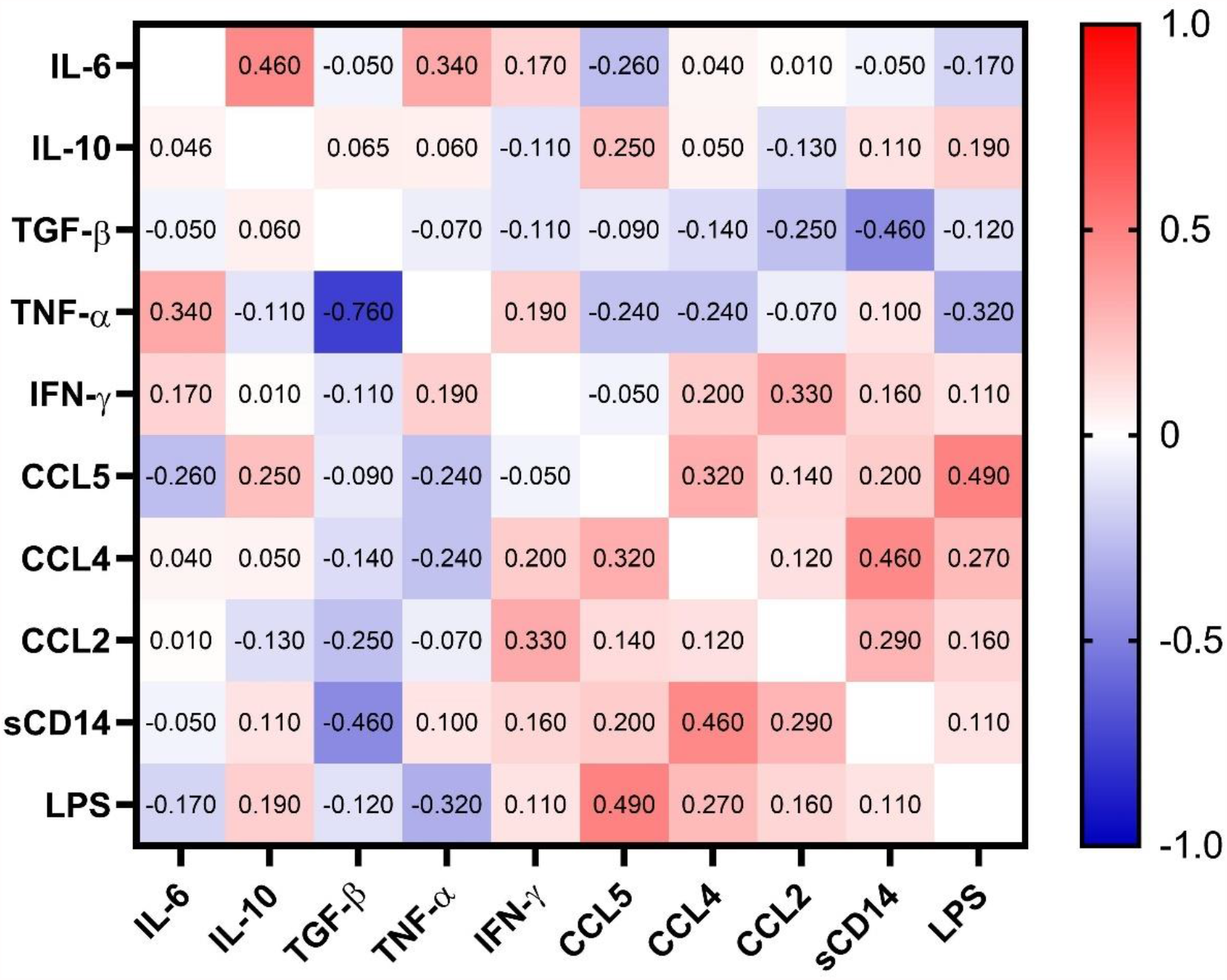
Correlation heat map including IL-6, IL-10, TGF-β, TNF-α, IFN-γ, CCL2/MCP-1, CCL4/MIP-1β, CCL5/RANTES, sCD14 and LPS, at admission to the Hospital. Pearson’s correlation coefficients are plotted. Cells were colored according to the strength and trend of correlations (shades of red=positive, shades of blue=negative correlations). Statistical analysis performed through Pearson’s coefficient correlation.

### Systemic inflammatory cytokines and chemokines and microbial translocation markers in COVID-19 survivors and non-survivors at T1 and T2

The time course of systemic inflammatory markers and variables related to microbial translocation in 14 survivors and 12 non-survivors COVID-19 patients were evaluated in two times. To this, peripheral blood of patients was collected at hospital admission (T1) and at the discharge time from the hospital (T2: 0 to 72 hours before leaving hospital or death). The time course of systemic inflammatory mediators was presented in Figure 3. Increases in IFN-γ (p<0.01) and TNF-α (p<0.05) concentrations were identified in survivors COVID-19 patients at T2. Non-survivors COVID-19 patients presented increased levels of IL-6 (p<0.01), TNF-α (p<0.05), CCL5/RANTES (p<0.05) and CCL4/MCP-1 (p<0.01) at T2.

**Figure 3.**
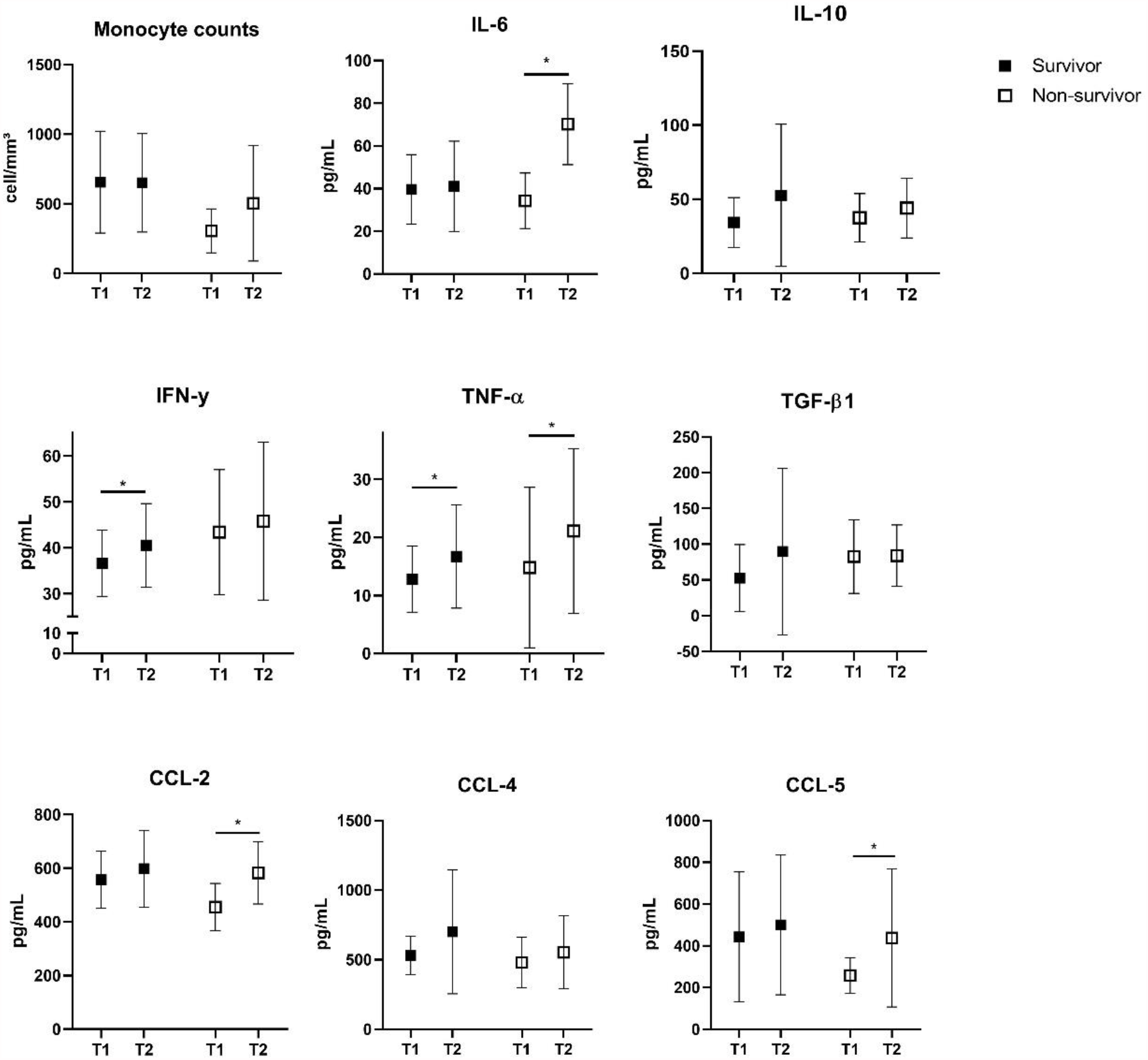
The systemic levels of IL-6, IL-10, IFN-γ, TNF-α, TGF-β1, CCL5/RANTES, CCL4/MIP-1β, and CCL2/MCP-1 in survivors and non-survivors COVID-19 patients at the hospital admission (T1) and at the end of hospitalization (T2). Data presented as mean ± SD. Two-way ANOVA followed by Bonferroni’s post hoc were applied to verify time and group effects (p<0.05). * Indicates p<0.05.

Non-survivor patients presented higher LPS levels in T2 compared to T1 (p<0.05), but sCD14 only tended to increase in T2 (p=0.055). On the other hand, no changes were identified in sCD14 and LPS at the end of hospitalization in COVID-19 survivors. Furthermore, the delta value (Δ, T2 – T1) of LPS levels positively correlated with Δ CCL2/MCP-1 levels (r = 0.54; p=0.008) (Figure 4). Together, these data indicate that the increase of LPS may be associated with death.

**Figure 4.**
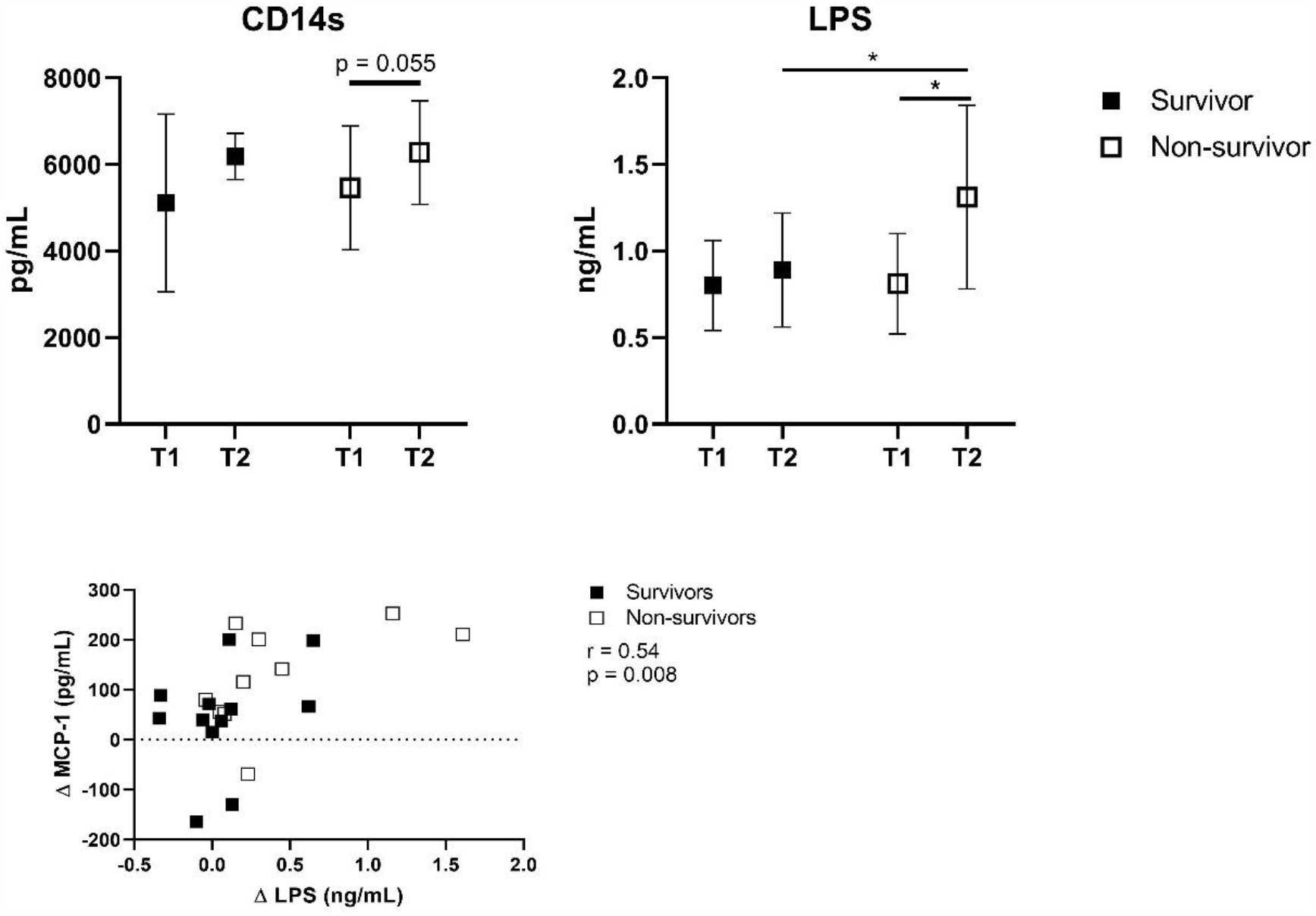
The systemic levels of soluble CD14 (A) and LPS (B) in survivors and non-survivors COVID-19 patients at the hospital admission (T1) and at the end of hospitalization (T2). Correlation between Δ MCP-1 and Δ LPS levels of COVID survivors and non-survivors are represented (C). Two-way ANOVA followed by Bonferroni’s post hoc were applied to verify time and group effects. (p<0.05). * Indicates p<0.05. Pearson’s Coefficient test were applied to verify the correlation between inflammatory markers and LPS levels (p<0.05).

### Effect of plasma from COVID survivors and non-survivors on THP-1 cell phenotype

As hospitalized non-survivors COVID-19 patients presented higher levels of microbial translocation markers with concomitant increase in some plasmatic inflammatory mediators, we hypothesized if the plasma from COVID-19 patients alters the phenotype of monocytes including subpopulation and activation markers and TLR4 and chemokine receptors expression. To this, we incubated THP-1 human monocyte lineage with plasma of survivors (n=8) and non-survivors (n=8) COVID-19 patients collected at hospital admission and at the end of hospitalization. Plasma incubation did not alter the cell viability in THP-1 cells evaluated by MTT assay (Suppl. Fig. 1). Figure 5 shows the expression of TLR4, CD14, CD16, CD69, HLA-DR, CCR2, CCR5 and CCR7 in THP-1 cells after incubation with LPS (1 ng/mL), and plasma from COVID-19 survivors or non-survivors patients collected at T1 and T2. Interestingly, plasma from COVID-19 non-survivor patients obtained at T2 increased the expression of TLR4 (p=0.03), CCR2 (p=0.05), CCR5 (p=0.02) and CCR7 (p=0.02) and the activation marker CD69 (p=0.02) on the cell surface of THP-1 cells compared to T1 plasma from the same group. The expression of CD16 (p=0.04) and CCR5 (p=0.03) were higher in THP-1 cells incubated with plasma from COVID-19 survivor patients collected at T2. No statistical differences were found in CD14 and HLA-DR expression in this study (p<0.05). These data indicate that non-survival COVID-19 patients presented higher levels of microbial translocation products and inflammatory mediators in the plasma that increase activation/migration status of monocytes.

**Figure 5.**
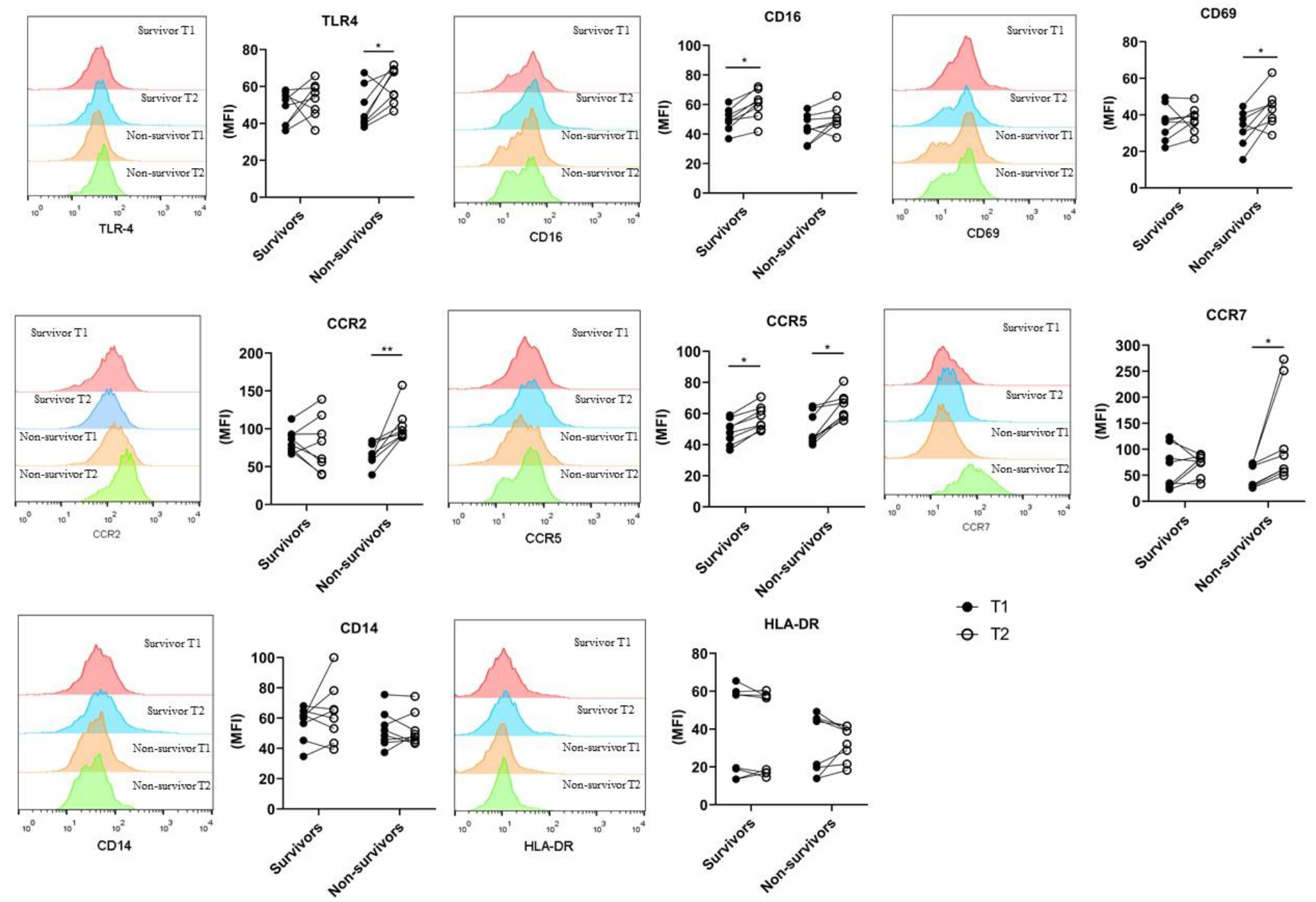
Effect of plasma from COVID survivors and non-survivors on THP-1 cell phenotype. THP-1 cells were incubated with plasma from COVID survivors and non-survivors at T1 and T2 for 15 h, then the expression of monocyte phenotype, activation markers and chemokine receptors were evaluated by flow cytometry. Media Fluorescence Intensity of T1 and T2 are represented. Two-way ANOVA followed by Bonferroni’s post hoc were applied to verify time and group effects. (p<0.05).

## Discussion

Hospitalized COVID-19 patients display a broad spectrum of inflammatory response, suggesting that systemic mediators contribute to the immune activation, disease severity and the outcome course in COVID-19 (20). In the present study, we hypothesized that microbial translocation markers could impact on inflammatory response in COVID-19 patients. Our main findings can be summarized as follows: a) markers of microbial translocation were higher at hospital admission in COVID-19 patients; b) COVID-19 patients presented a proinflammatory status as viewed by the higher levels of IL-6, IFN-γ, TNF-α, CCL5/RANTES, CCL4/MIP-1β and CCL2/MCP-1; c) non-survivors COVID-19 patients presented increases in LPS and sCD14 during the hospitalization associated with increased IL-6, TNF-α, CCL5/RANTES and CCL2/MCP-1 systemic levels; d) survivors COVID-19 patients maintained LPS and sCD14 levels at stable values concomitant with increases in IFN-γ levels; e) the incubation of non-survivors COVID-19 plasma obtained in the T2 hospitalization increased the expression of TLR4, CCR2, CCR5, and CD69 on the cell surface of THP-1 monocytic lineage, while THP-1 cells incubated with T2 plasma of survivor patients presented higher CCR5 and CD16 expression. In conjunct, our data indicates that increased microbial translocation during hospitalization coexist with the inflammatory condition of SARS-CoV-2 infection and could lead to the higher monocyte activation which may be associated with worsening of COVID-19 outcomes, such as death.

Angiotensin-Converting Enzyme 2 (ACE-2), the most recognized receptor that binds with the Spike protein of SARS-CoV-2 and allows the host infection, is widely expressed in epithelial cells along the gastrointestinal (GI) tract (8,21). Thus, SARS-CoV-2 can invade the intestinal tract causing GI dysfunction which disrupt intestinal barriers, increasing the translocation of microbial products (9,21). Increased systemic endotoxemia has been previously described in several pathological conditions associated with hospitalization, becoming a recognized marker of critical ill who suffer sepsis (22–24). Furthermore, recent studies described disrupted gut barrier integrity and increased circulating bacteriome in moderate and severe COVID-19 patients (9,16,17). Here, we found increased LPS and sCD14 levels in the plasma of hospitalized COVID-19 patients at admission and during ICU hospitalization. Arunachalam and colleagues (20) found higher levels of bacterial DNA products, as measured by PCR quantification of 16S ribosomal RNA gene product, and LPS levels in severe ICU patients. The increased bacterial products in the peripheral circulation may contribute to the enhanced macrophage activation in infected tissue (i.e. lungs and gastrointestinal tissues) and to the failure to suppress the hyperinflammation during SARS-CoV-2 infection. In fact, elevated markers level of gut leakage was previously associated with inflammasome activation which leads to cardiac injury during the course of hospitalization in COVID-19 patients (17). Corroborating these data, other studies show an important role for endotoxin and sCD14 in heart failure (25,26).

Microbial translocation to the peripheral blood also contributes to an early activation of monocytic cells. Indeed, we observed higher sCD14 in survivors and non-survivors COVID-19 patients. In line with this, Bowman and coworkers (27) found increased sCD14 levels in COVID-19 patients independent of the disease severity degree. Furthermore, the authors revealed that COVID-19 critical ill patients who recovered or deceased had the highest sCD14 and lipopolysaccharide-binding protein (LBP, another marker of endotoxemia) values compared to the other groups (27). These data underlined the presence of strong activation of the monocytic lineage on hospital admission due to SARS-CoV-2 infection in conjunct with LPS translocation and inflammatory exacerbation in ICU patients (28). In according with this study, systemic markers of monocyte activation, mainly sCD14 and sCD163, correlated with inflammatory cytokines in a non-intensive care unit COVID-19 cohort patients (29).

It was previously postulated that perturbations in gut microbiota and microbial translocation may exacerbate the severity of cytokine storm in COVID-19 (30). Moreover, sCD14 tended to be higher at T2 in non-survivors COVID-19 patients. Interestingly, heat map correlations revealed that sCD14 correlated with CCL4/MIP-1β and CCL2/MCP-1, while LPS correlated with CCL5/RANTES and CCL4/MIP-1β. These results suggest that microbial translocation may contribute to the increases in chemokines, which recruits innate immune cells to the infectious site. In fact, chemokines release is directly involved in the acute respiratory disease syndrome, a major complication leading to death in severe COVID-19 infection (31,32). All the chemokines evaluated here, CCL4/MIP-1β, CCL2/MCP-1 and CCL5/RANTES, are upregulated early post SARS-CoV-2 infection and contribute to the migration of inflammatory monocytes, natural killer cells and T cells to the infection site (33). Thus, chemokines seem to be vital players during COVID-19 pathogenesis and the hyperinflammatory condition of severe disease. Our data also indicate that there was no difference between COVID-19 groups at hospital admission regarding chemokine or cytokine concentrations.

However, we found increases in the systemic LPS levels concomitant with higher IL-6, TNF-α, CCL/5/RANTES and CCL2/MCP-1 concentrations in the plasma of non-survivors group collected 24-72 h before the death of critical COVID-19 patients. Lucas and coworkers (4) described a deregulated increase in inflammatory cytokines and chemokines in severe COVID-19 illness patients who deceased during the hospitalization, suggesting a maladapted immune response, and exacerbated inflammatory induction associated with severe COVID-19 and poor clinical outcome. Interestingly, COVID-19 admitted to intensive care units have strong cytokine response compared to other types of pneumonia, and elevations of IL-6 levels during hospitalization predicts ICU stay and mortality (34). Thus, the increases of inflammatory mediators occur in conjunct with increased microbial translocations, as suggested by increased LPS and sCD14 levels, in the blood of COVID-19 deceased patients.

The enhanced hyperinflammatory condition associated with higher LPS levels at T2 in non-survivors COVID-19 patients may contribute to systemic organ dysfunction. In this line, gut permeability may increase during the hospitalization course and contribute to the worsening of clinical outcomes because of respiratory distress syndrome caused by SARS-CoV-2 infection. Bacterial translocation during COVID-19 hospitalization strongly correlates with immune activation, increased proinflammatory production by innate immune cells and higher mortality rate (35). However, the interplay between systemic inflammation and LPS release in the blood remains to be elucidated in future studies involving COVID-19 cohorts.

Here, we hypothesized that serum factors, such as LPS and inflammatory cytokines and chemokines, may directly impact the phenotype of monocytes. To address this question, we incubated monocytic THP-1 lineage with the plasma samples of hospitalized survivors and non-survivors COVID-19 patients obtained at hospital admission and in a time compromising 0-72 hours before the final event (T1 and T2, respectively). Interestingly, TLR4, the LPS ligand, increased only in non-survivors COVID-19 patients which corroborates with the higher systemic LPS levels in this group. In fact, critically ill COVID-19 patients presented upregulation in TLR4 expression and the disruption in pro-inflammatory cytokine and chemokine production after monocyte TLR4 stimulation similarly to the observed in bacterial sepsis (36).

Furthermore, spike protein binds to LPS to activate TLR4/NF-kB axis to modulate the phenotype of monocytes. In this sense, we found increased CD69 expression on THP-1 cells incubated with T2 plasma of non-survivors COVID-19 patients, indicating an early monocyte activation. However, no difference was observed in HLA-DR expression on THP-1 cells after incubation with plasma of both groups. Thus, plasma obtained from COVID-19 patients may have no impact on antigen presentation and long-term activation of monocytes. On the other hand, plasma from survivors COVID-19 patients collected at T2 moment increased CD16 expression in THP-1 cells. Data from literature described higher percentages of CD14^+^CD16^high^ monocytes and very low expression of HLA-DR in all CD14^+^ monocyte subsets in severe COVID-19 patients (37–39). In this sense, the mobilization of newly bone marrow monocytes associated with continuous higher systemic and organ inflammation along the course of the acute infection, rather than short-term stimulation, may impact in the changes of CD16 and HLA-DR expression in circulating monocytes.

Severe COVID-19 patients presented an inflammatory phenotype of monocytes associated with early activation and downregulation of HLA-DR (38,40). Monocytes upregulate CD69 expression which promotes tissue infiltration and retention(41,42). Increased activation and CD69 expression in monocytes of severe COVID-19 subjects was associated with diapedesis and tissue infiltration, contributing to the monocytopenia observed in the peripheral circulation of diseased patients(43). In this line, the increased expression of CCR2, CCR5 and CCR7 in THP-1 incubated with T2 plasma of non-survivors COVID-19 patients may indicate the migratory stimulation of soluble factors during SARS-CoV-2 infection. Moreover, THP-1 cells also presented higher CCR5 expression after the incubation with plasma of survivors COVID-19 patients obtained at T2. Infiltrating monocytes constitute the majority of leukocytes migrating into the infected lungs, contributing to the severe lung inflammation in COVID-19 (40). In fact, increased transcription of CCR2 and CCR5 in macrophages present in bronchial alveolar lavage fluid of severe COVID-19 patients (44). Furthermore, increased CCR2^+^ monocytes elicit calcium influx, producing higher levels of reactive oxygen radicals and upregulates integrin expression to recruit neutrophils and mast cells, which exacerbates the inflammatory local response and induces tissue damage (45–47). The activation of CCR5^+^ in macrophages is linked to the signal transduction downstream of Gαi/PI3K/AKT and Gαi/MEK/ERK pathways that induce anti-apoptotic activity, maintaining inflammatory exhausted macrophages in the inflamed lung (48). Interestingly, COVID-19 patients treated with lerolimab, an antibody used to block CCR5, presented a rapid reduction of plasma IL-6, restoration of the CD4/CD8 ratio, and a significant decrease in SARS-CoV-2 plasma viremia (49). Antigen uptake by human monocyte derived dendritic cells, during viral infections, increases the expression of CCR7 that mediates the migration of antigen-bearing cells to lymphatic tissue (50). Thus, the upregulation of CCR7 is crucial for the recruitment of antigen presenting cells to secondary lymphoid organs during viral infections (51). Due to their pivotal role in the migration of inflammatory monocytes to the infected tissues and lymphoid organs, future experimental studies should be conducted to evaluate the impact of monoclonal antibody therapies that blockade the overexpression of chemokine receptors in severe COVID-19 hospitalization.

In conclusion, this study shed light on the modulation of microbial translocation markers associated with inflammatory cytokines in the peripheral blood of survivors and non-survivors COVID-19 patients during hospitalization. We also noted that plasma samples of non-survivors COVID-19 patients impacts on the phenotype of monocyte lineage cells, increasing the expression of early activation markers and selected chemokine receptors. Our data indicates that deceased patients present a condition characterized by microbial translocation that coexist with the inflammatory condition of SARS-CoV-2 infection and could contribute to the observed alterations in the monocyte phenotype and function.

## Data Availability

Data will be available when requested.

